# Differential Associations of Postoperative Plasma and Cerebrospinal Fluid Albumin Changes with Delirium Following Non-Cardiac Surgery

**DOI:** 10.64898/2026.09.18.26363418

**Authors:** Refaat Hassan, Ligia S. Dumont, Mary Cooter Wright, Jeffry Takla, Kristen Monten, Samuel Teshome, Edward R. Marcantonio, Niccolò Terrando, Jeffrey N. Browndyke, Heather E. Whitson, Harvey J. Cohen, Andrea G. Nackley, Megan K. Wong, Marguerita E. Klein, Piper C. Boykin, Noah J. Timko, Michael Muehlbauer, E. Wesley Ely, Joseph P. Mathew, Miles Berger, Michael J. Devinney, the MADCO-PC, INTUIT Study Teams

## Abstract

**BACKGROUND:** Postoperative increases in cerebrospinal fluid (CSF) to plasma albumin ratio (CPAR), a blood-brain barrier dysfunction marker, have been associated with postoperative delirium (POD) and prolonged hospital stay. However, the contributions of plasma versus CSF albumin changes remain unclear.

**AIM:** To determine whether 24-hour postoperative changes in plasma and CSF albumin are independently associated with POD and hospital length of stay.

**METHODS:** Plasma and CSF albumin concentrations were measured before and 24-hours postoperatively in older non-cardiac surgery patients. Multivariable logistic and negative binomial regression models evaluated associations between albumin changes and POD or hospital length of stay, adjusting for age, baseline cognition, surgery type, and fluid administration.

**RESULTS:** Of 240 patients, 31 (13.3%) developed POD. From before to 24-hours after surgery, both plasma albumin (median 3.95 to 3.43 g/dL, p<0.001) and CSF albumin decreased (median 22.68 to 20.43 mg/dL, p<0.001). Greater plasma albumin decreases (per 0.43 g/dL) were independently associated with POD (OR 2.14, 95% CI 1.28-3.58; p=0.004) and longer hospital stay (mean ratio 1.44, 95% CI 1.26-1.64; p<0.001). In contrast, CSF albumin changes (per 5.8 mg/dL higher) were not associated with POD (OR 0.97, 95% CI 0.60-1.58; p=0.91) but were modestly associated with longer hospital stay (mean ratio 1.17, 95% CI 1.04-1.32; p=0.01).

**CONCLUSIONS:** Plasma and CSF albumin decreased postoperatively, but only plasma decreases were associated with POD. These results suggest that associations between CPAR increases and POD may be due to postoperative plasma albumin decreases and highlight limitations of CPAR as a standalone marker of postoperative BBB dysfunction.

## Introduction

Postoperative delirium is a common acute neurocognitive complication affecting 15– 50% of older surgical patients,^1^ which is associated with longer hospital stays, over $32 billion in annual US healthcare costs, and increased mortality risk.^2, 3^ Despite its prevalence and impact, the pathophysiological mechanisms of postoperative delirium remain incompletely understood.

One potential mechanism of postoperative delirium is blood-brain barrier (BBB) dysfunction. In mice, surgery induces BBB disruption that is associated with subsequent attentional and cognitive deficits.^4^ In humans, postoperative delirium has been associated with higher plasma S100β,^5^ a non-specific BBB marker, and with greater postoperative increases in cerebrospinal fluid (CSF) to plasma albumin ratio (CPAR),^5, 6^ an established marker of CNS barrier permeability.^7^ While these studies support BBB dysfunction as a plausible contributor to delirium, human biomarkers have important limitations.

Interpretation of postoperative CPAR is complicated by substantial postoperative decreases in plasma albumin, as plasma albumin decreases by approximately 30% just 1 day after major non-cardiac surgery.^8^ Kinetic studies of albumin equilibration between plasma and CSF suggest that complete equilibration requires approximately 24-72 hours.^9^ Thus, rapid postoperative plasma albumin decreases could temporarily outpace corresponding changes in CSF albumin, such that 24-hour postoperative CPAR increases could reflect transient disequilibrium between compartments rather than increased transfer of albumin into the CSF.^10^ Consequently, postoperative CPAR increases may be influenced by rapid decreases in plasma albumin concentrations rather than solely by changes in BBB permeability. However, prior studies have not examined paired acute postoperative changes in plasma and CSF albumin to determine how changes in each compartment contribute to postoperative CPAR increases and delirium.

Acute postoperative plasma albumin decreases may also be informative of delirium risk independent of their effect on CPAR. Postoperative hypoalbuminemia has been associated with delirium,^11^ acute kidney injury,^12^ and mortality.^13^ Hypoalbuminemia likely serves as a marker of other pathophysiological processes contributing to these outcomes rather than a direct cause, because randomized trials of albumin-containing fluids versus crystalloids have not consistently improved outcomes in surgical patients.^14^ Furthermore, the early postoperative decrease in plasma albumin likely reflects multiple processes, particularly inflammation-induced increases in capillary permeability and redistribution of albumin into the extravascular space,^15, 16^ together with perioperative hemodilution.^8^ Although albumin is classified as a negative acute-phase reactant (i.e., decreases during inflammation), reduced hepatic synthesis alone is unlikely to account for the rapid and significant decrease observed within 24 hours, especially since albumin synthesis is maintained or increased after surgery.^15,16^ Therefore, postoperative plasma albumin decreases may serve as a marker of delirium risk because it reflects multiple perioperative insults, such as inflammation, endothelial injury, and fluid shifts.

Since postoperative plasma albumin decreases reflect multiple physiological disturbances, acute postoperative plasma albumin decreases may also be associated with proxy measures of overall postoperative recovery, such as postoperative hospital length of stay.^17^ Although prolonged hospitalisation occurs in patients with delirium,^6, 18^ it is possible that postoperative plasma albumin changes may be independently associated with both delirium risk and overall postoperative recovery.

We hypothesized that postoperative changes in plasma and CSF albumin would contribute differently to postoperative CPAR increases and that postoperative plasma albumin decreases would be more strongly associated with postoperative delirium and hospital length of stay than postoperative CSF albumin changes. To test these hypotheses, we carried out a secondary analysis of plasma and CSF albumin levels from our prior study that examined the association between postoperative CPAR changes and postoperative delirium.^6^

## Methods

### Study Information

Data and biofluid samples are from two prospective cohort studies (see study timeline: Figure S1) for: Markers of Alzheimer’s Disease (AD) and Neurocognitive Outcomes after Perioperative Care (MADCO-PC; NCT01993836)^19^ and Investigating Neuroinflammation Underlying Post-operative Cognitive Dysfunction (INTUIT; NCT03273335).^20^ Both studies were approved by the Duke Institutional Review Board, and all participants provided written informed consent prior to enrolment.^19, 20^ Eligible patients were aged ≥60 years undergoing non-cardiac, non-neurologic surgery scheduled for ≥2 hours.^19, 20^ Exclusion criteria included chemotherapy (due to potential cognitive side effects) or inability to undergo lumbar puncture. INTUIT additionally excluded patients on anti-inflammatory or immunomodulatory drugs before surgery.

### CSF and Blood Collection, Processing and Albumin Assays

CSF and blood samples were collected preoperatively and 24-hours postoperatively and processed within one hour as previously described for storage at -80°C. CSF and plasma albumin levels were measured in duplicate by technicians blinded to delirium status or other patient factors (see Supplementary Materials for full details).^6^ Bromocresol purple dye binding and an immunoturbidimetric microalbumin assay on the UniCel DxC 600 System (Beckman Coulter) was used to measure plasma (CV 0.74%) and CSF (CV 1.2%) albumin, respectively.

### Cognitive Testing

Preoperative cognitive function was assessed with a 14-item test battery used in prior studies of postoperative cognitive dysfunction,^21^ Cognition was summarised by global and domain scores as described (see Supplementary Materials).^6, 21^

### Delirium Assessment

Delirium was assessed daily in MADCO-PC with the Confusion Assessment Method (CAM) using a skip pattern (i.e., each CAM was terminated if the patient was not inattentive, since inattention must be present for a patient to have delirium), and twice daily until day 5 postoperatively using the 3-minute Diagnostic CAM (3D-CAM) in INTUIT.^20^ The Confusion Assessment Method for the Intensive Care Unit (CAM-ICU) was employed in both studies to assess delirium in intubated or otherwise nonverbal patients.^22^ Additionally, a validated chart review method was utilized to identify any delirium cases that may have been missed due to symptom fluctuation.^23^ Postoperative delirium was defined as at least one positive CAM assessment (3D-CAM, CAM, or CAM-ICU) or a positive chart review for delirium.

### Data Collection

Subject characteristics and clinical variables (such as hospital length of stay, crystalloid and albumin-containing solution administration, blood product utilisation) were extracted from the electronic medical record and study records. Charlson comorbidity scores were determined and *APOE4* genotyping was performed as described.^6^ The Duke Activity Status Questionnaire was administered preoperatively.^24^

### Data Analysis

Sample size was selected based on our prior work in which >200 patients provided >80% power to detect relationships between changes in CSF-to-plasma albumin ratios (as a continuous measure) and postoperative delirium.^6^ Preoperative-to-24-hour postoperative changes in plasma and CSF albumin were examined with Wilcoxon signed rank tests. Wilcoxon rank sum tests and univariable logistic regression were used to evaluate the relationships between postoperative delirium and both plasma and CSF albumin levels. Multivariable negative binomial regression analysed associations between postoperative changes in plasma or CSF albumin levels and postoperative length of stay. All multivariable models were adjusted for age, baseline cognitive function (both of which are previously identified delirium risk factors)^3^ and surgery type (given that patients undergoing different types of surgery often differ in baseline characteristics). To study the association of postoperative albumin levels with outcomes of interest, models included adjustment terms for preoperative albumin levels to account for baseline albumin status (in either plasma or CSF) and 24-hour perioperative fluid administration (albumin and crystalloid per ideal body weight) to account for dilutional (crystalloids) or iatrogenic (exogenous albumin) effects on albumin levels. We did not include CPAR together with plasma and CSF albumin in the same regression models because CPAR is calculated directly from plasma and CSF albumin concentrations. Inclusion of CPAR alongside either of its component variables would violate model independence assumptions and could lead to unstable coefficient estimates due to multicollinearity. Delirium occurrence was included as an adjustment term for postoperative length of stay models.

We report odds ratio (ORs) and 95% confidence intervals (CIs) from regression models for delirium, and mean ratio (MRs) and incidence rate ratios (IRRs) with 95% CIs from negative binomial regression models for postoperative length of stay. We report effects for a 1 SD decrease for plasma albumin, and for a 1 SD increase in CSF albumin. Statistical analysis was conducted in SAS (SAS INC, Cary, NC; v 9.4) and R (R Foundation for Statistical Computing, Vienna, Austria; v4.4).^25^ Significance was set at α = 0.05; all tests were two sided.

## Results

### Subject Characteristics

Subject enrolment is detailed in Figure 1, and baseline characteristics are presented in Table 1. Median [Q1, Q3] estimated blood loss (EBL) was 50 mL [10, 200], 5/240 (2.1%) patients received red blood cell transfusions, and no patients received fresh frozen plasma, platelets, or cryoprecipitate. Compared to those without delirium, patients who developed postoperative delirium had fewer years of education, lower baseline continuous cognitive index scores (a sensitive global measure of cognition)^26^ and Mini-Mental State Examination scores. Of the 31 delirium cases, 28 cases were hypoactive, 2 hyperactive, and 1 mixed. Median duration of delirium was 1-day (26 cases had 1-day duration), and all delirium cases resolved before discharge.

**Figure 1:**
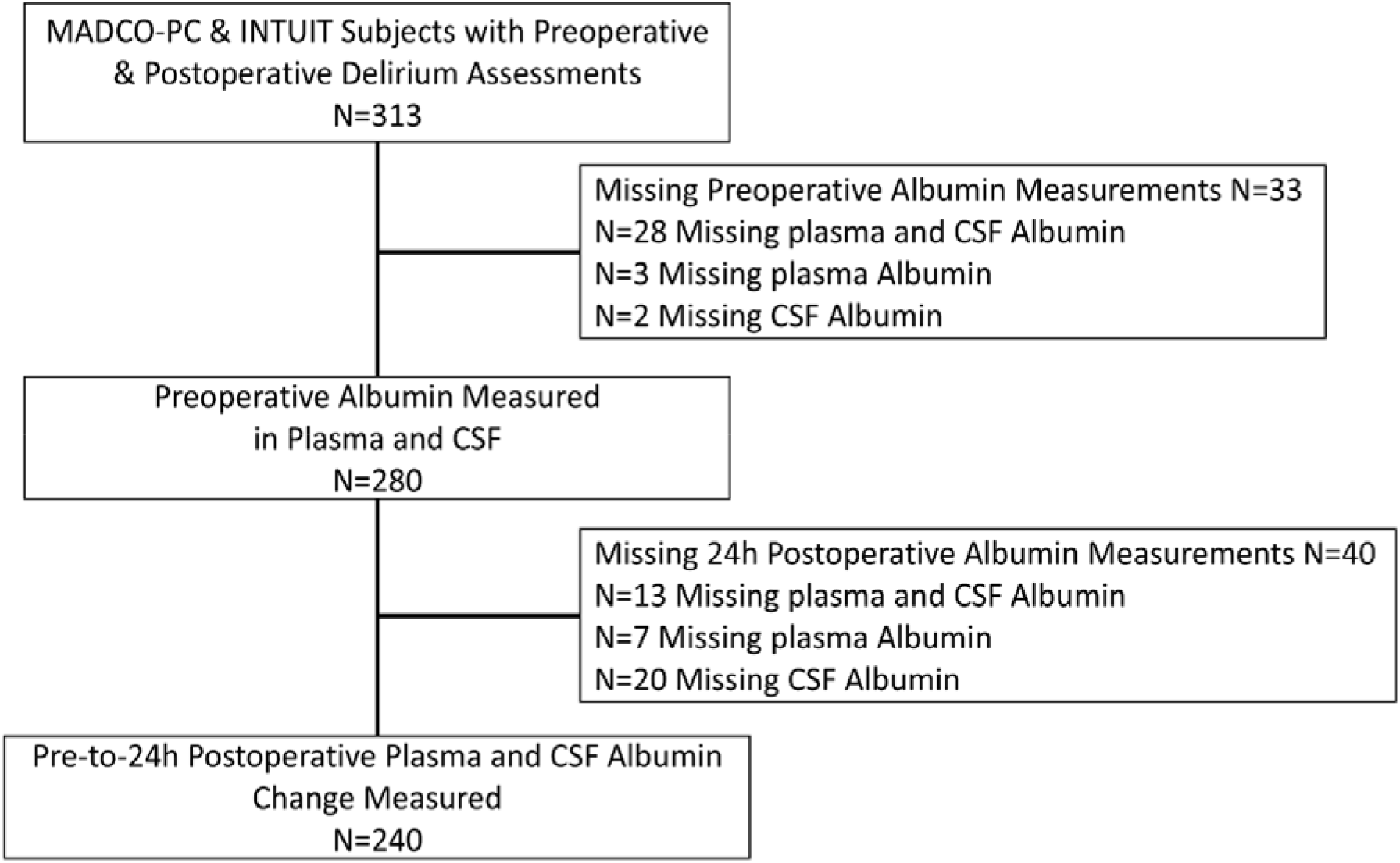
Subject enrolment flow diagram.

**Table 1:**
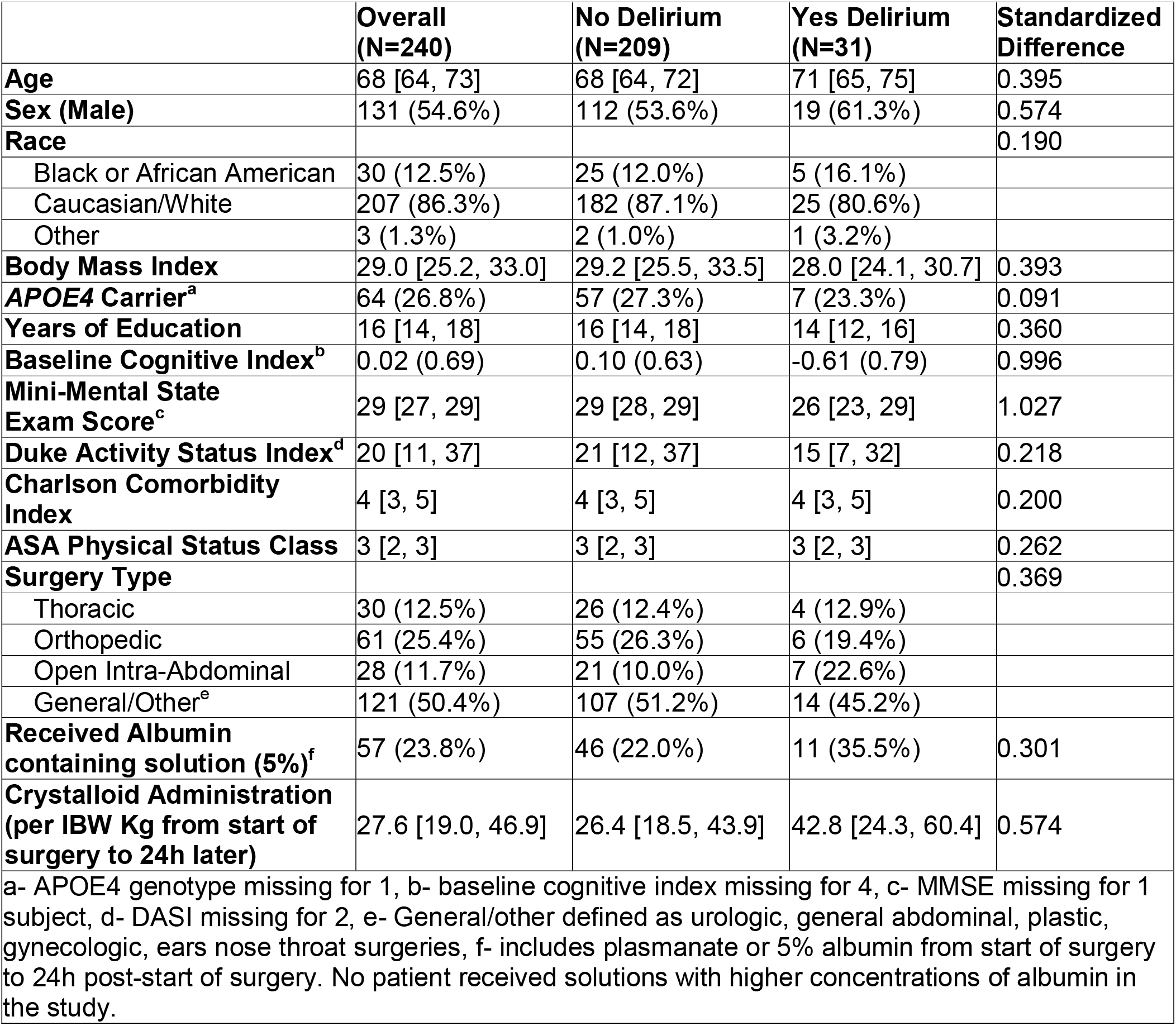
Subject characteristics.

### Postoperative Changes in Albumin Levels

There were significant decreases in plasma albumin from before surgery (median [Q1, Q3] 3.95 g/dL [3.70, 4.20]) to 24-hour postoperative (3.43 g/dL [3.10, 3.75], p<0.001) and in CSF albumin levels before surgery (22.68 mg/dL [16.70, 28.10]) to 24-hour postoperative (20.43 mg/dL [15.0, 27.45], p<0.001; Figure S2). CPAR change was negatively correlated with preoperative-to-24-hour postoperative plasma albumin change (Figure S3A, rho=−0.48, 95% CI -0.57, -0.38; p<0.001), and also positively correlated with CSF albumin change (Figure S3B, rho=0.81, 95% CI 0.76, 0.85; p<0.001).

### Albumin Levels and Postoperative Delirium

Preoperative plasma albumin levels did not differ between groups (Table 2, Figure 2A). However, 24-hour postoperative plasma levels were lower in patients who developed versus those who did not develop delirium (Table 2, Figure 2B). There were larger preoperative-to-24-hour postoperative plasma albumin decreases in patients who developed versus those who did not develop postoperative delirium (Table 2, Figure 2C).

**Figure 2:**
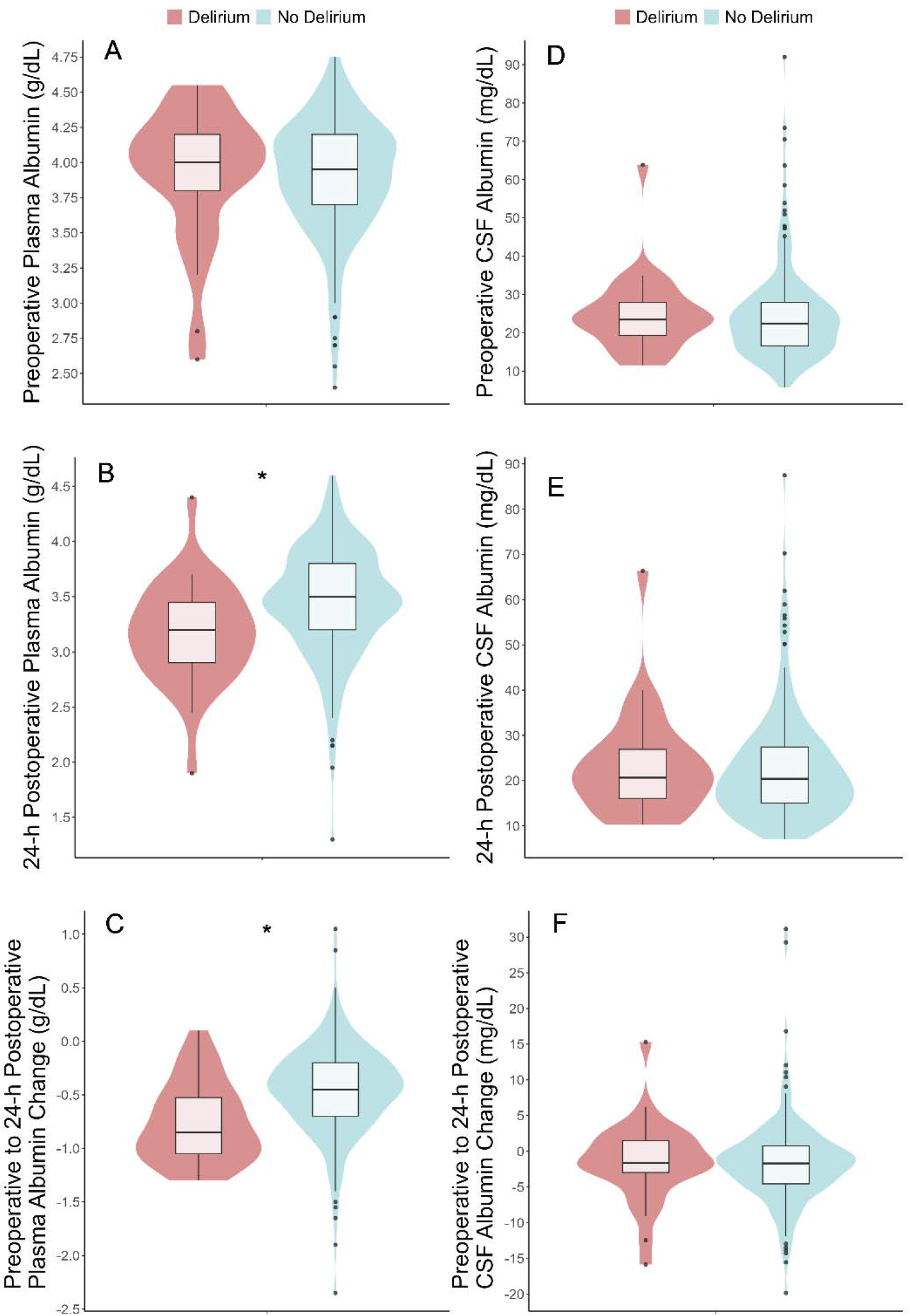
Comparison of preoperative and postoperative plasma and CSF albumin levels in patients with versus without postoperative delirium. (A) Preoperative plasma albumin levels, (B) 24-hour postoperative plasma albumin levels, (C) preoperative-to-24-hour postoperative plasma albumin levels (D) preoperative CSF albumin levels, (E) 24-hour postoperative CSF albumin levels, (F) preoperative-to-24-hour postoperative CSF albumin levels, in those with (n=31) versus those without (n=209) postoperative delirium. *p<0.001 in Wilcoxon rank sum test

**Table 2:**
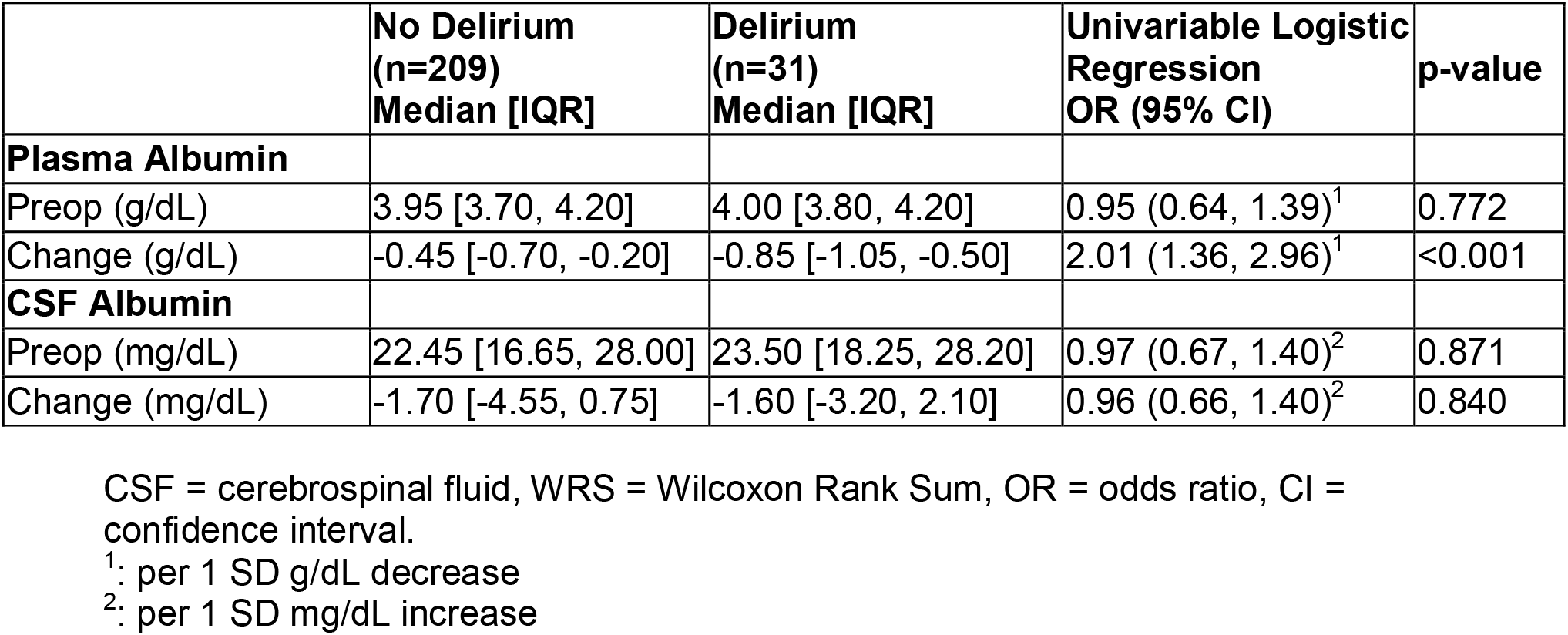
Univariable associations between plasma albumin and CSF albumin, and postoperative delirium.

|  | No Delirium<br>(n=209)<br>Median [IQR] | Delirium<br>(n=31)<br>Median [IQR] | Univariable Logistic<br>Regression<br>OR (95% CI) | p-value |
| --- | --- | --- | --- | --- |
| **Plasma Albumin** |  |  |  |  |
| Preop (g/dL) | 3.95 [3.70, 4.20] | 4.00 [3.80, 4.20] | 0.95 (0.64, 1.39)<sup>1</sup> | 0.772 |
| Change (g/dL) | -0.45 [-0.70, -0.20] | -0.85 [-1.05, -0.50] | 2.01 (1.36, 2.96)<sup>1</sup> | <0.001 |
| **CSF Albumin** |  |  |  |  |
| Preop (mg/dL) | 22.45 [16.65, 28.00] | 23.50 [18.25, 28.20] | 0.97 (0.67, 1.40)<sup>2</sup> | 0.871 |
| Change (mg/dL) | -1.70 [-4.55, 0.75] | -1.60 [-3.20, 2.10] | 0.96 (0.66, 1.40)<sup>2</sup> | 0.840 |

CSF = cerebrospinal fluid, WRS = Wilcoxon Rank Sum, OR = odds ratio, CI = confidence interval.
<sup>1</sup>: per 1 SD g/dL decrease
<sup>2</sup>: per 1 SD mg/dL increase

Preoperative plasma albumin levels were not associated with delirium in either univariable (OR=0.95 per 0.42 g/dL, 95% CI 0.64-1.39, p=0.78, Table 2) or multivariable analyses (Table S1). In a multivariable logistic regression adjusted for albumin and crystalloid administration (per kg ideal body weight), baseline plasma albumin, baseline cognitive function (continuous cognitive index), age, and surgery type, greater preoperative-to-24-hour postoperative plasma albumin decreases (OR=2.09 per 0.43 g/dL, 95% CI 1.25-3.49, p=0.005, Table S2) were associated with delirium. There was an additive effect of preoperative-to-24-hour postoperative plasma albumin change and baseline global cognitive function on delirium probability, meaning those with larger preoperative-to-24-hour postoperative plasma albumin decreases and lower baseline global cognitive function were at highest risk of delirium (Figure 3A). Interestingly, we also observed an independent association of volume of crystalloid administration with postoperative delirium (Table S2). In a sensitivity analysis that excludes patients who were delirious on or before postoperative day 1 (n=16 with onset of delirium after POD1), greater preoperative-to-24-hour postoperative plasma albumin decreases were still associated with postoperative delirium in both univariable (OR=1.84 per 0.43 g/dL, 95% CI 1.14-2.98, p=0.013) and multivariable analyses (OR=2.13 per 0.43 g/dL, 95% CI 1.21-3.75, p=0.009; Table S3).

**Figure 3:**
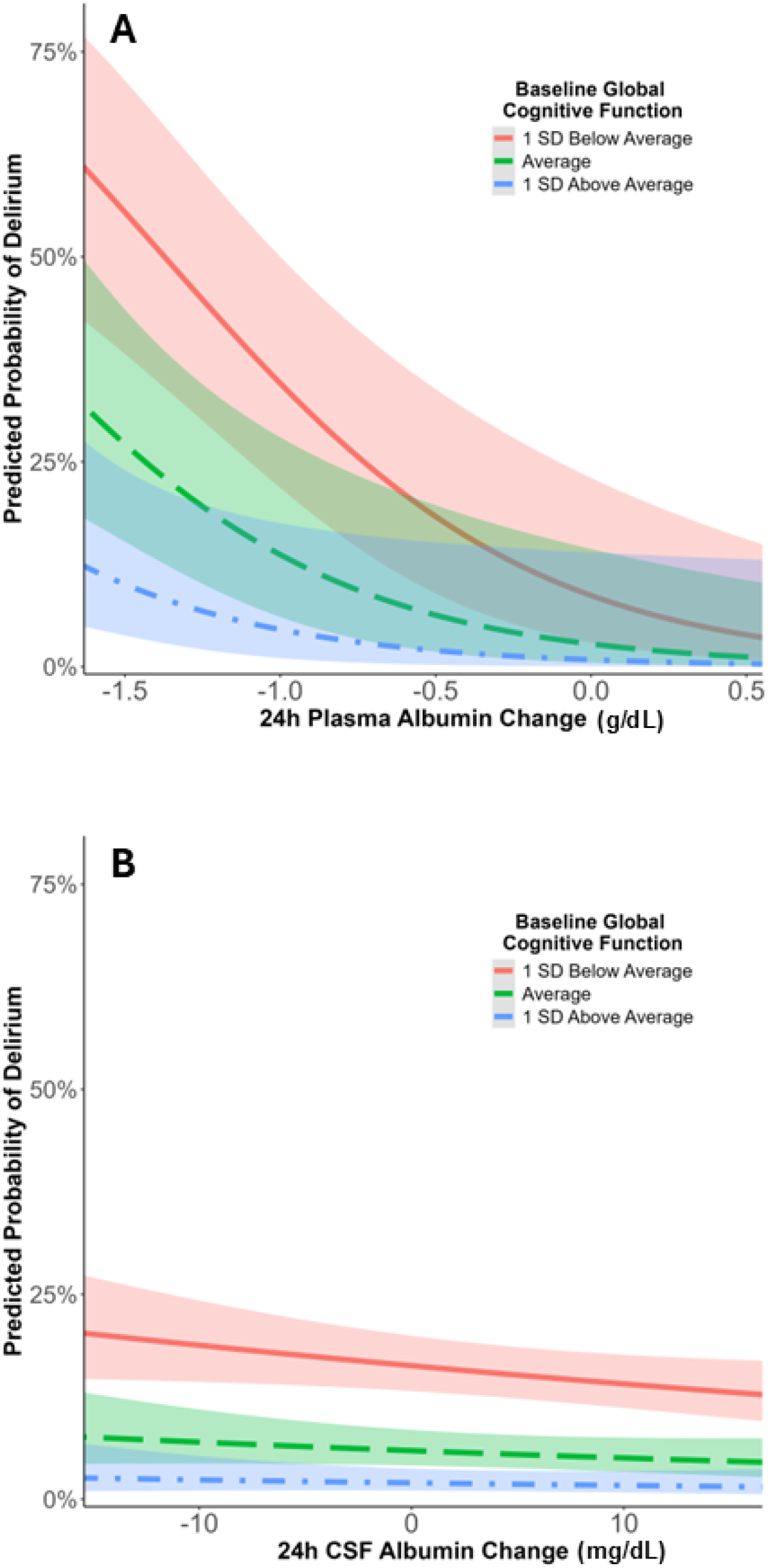
Predicted probability of delirium based on preoperative-to-24-hour postoperative (A) plasma albumin change and (B) CSF albumin change. (A) Predicted probability of postoperative delirium over the observed range of preoperative to 24-hour postoperative plasma albumin according to baseline global cognitive function in a multivariable logistic regression model adjusting for age, baseline global cognitive function, surgery type, baseline plasma albumin and intraoperative-to-24-hour postoperative intravenous albumin and crystalloid administration. (B) Predicted probability of postoperative delirium over the observed range of preoperative-to-24-hour postoperative CSF albumin according to baseline global cognitive function in a multivariable logistic regression model adjusting for age, baseline cognitive function, surgery type, baseline CSF albumin and intraoperative-to-24-hour postoperative intravenous albumin and crystalloid administration. Shaded areas represent mean prediction error.

**Figure 4:**
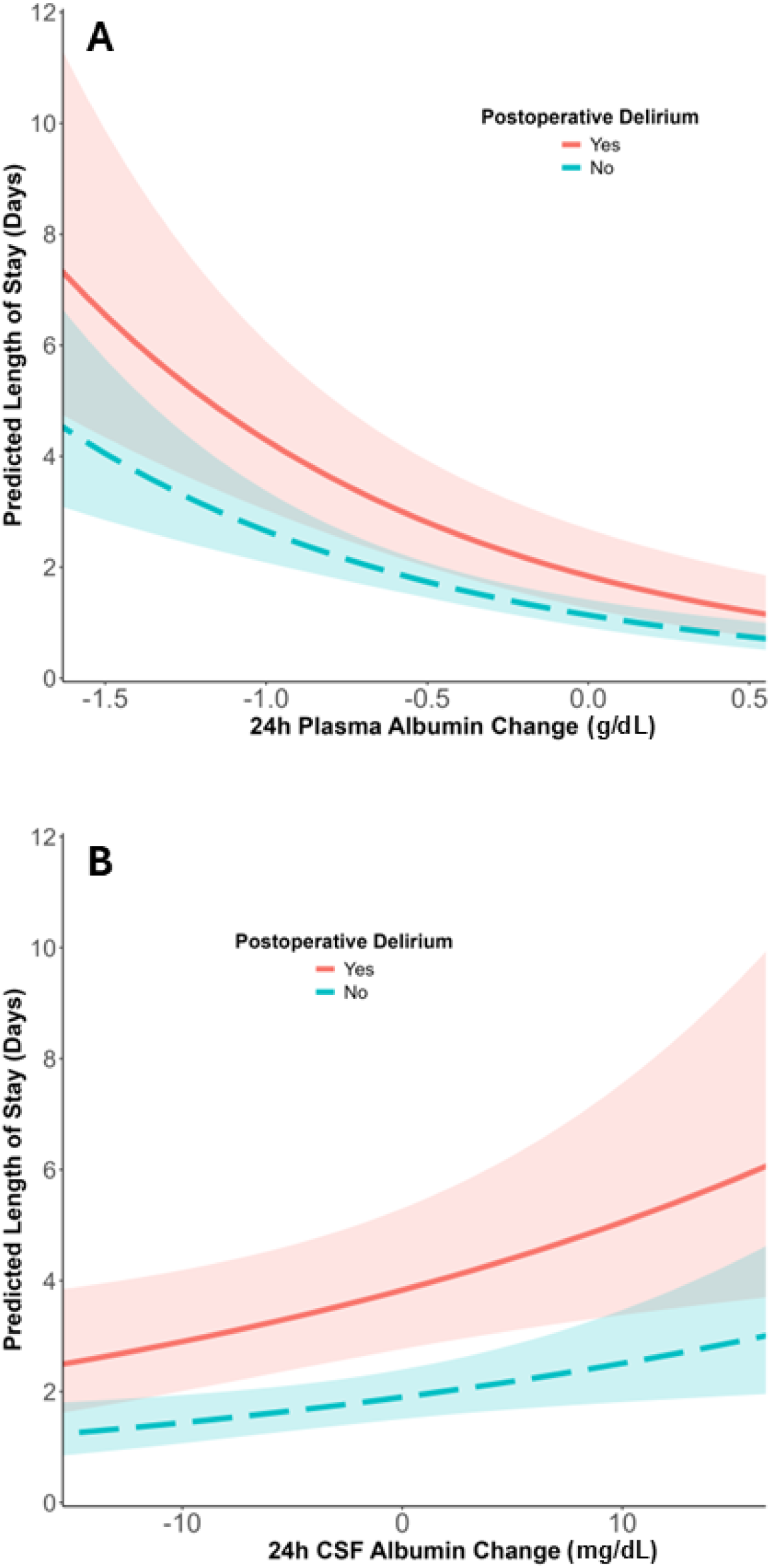
Predicted length of hospital stay based on preoperative-to-24-hour postoperative (A) plasma albumin change and (B) CSF albumin change. (A) Predicted length of postoperative hospital length of stay over the observed range of preoperative-to-24-hour postoperative plasma albumin change according to the presence or absence of postoperative delirium, in a multivariable negative binomial regression model adjusted for baseline plasma albumin, baseline global cognitive function, age, surgery type and intraoperative-to-24-hour postoperative albumin and crystalloid administration. Shaded areas represent mean prediction error. (B) Predicted length of postoperative hospital length of stay over the observed range of preoperative-to-24-hour postoperative plasma albumin change according to the presence or absence of postoperative delirium, in a multivariable negative binomial regression model adjusting for baseline CSF albumin, baseline global cognitive function, age, surgery type and intraoperative-to-24-hour postoperative albumin and crystalloid administration. Shaded areas represent mean prediction error.

Preoperative CSF albumin levels were not associated with postoperative delirium in univariable (Table 2) or multivariable logistic regression adjusting for baseline cognition, age, and surgery type (OR 1.09 per 11.6 mg/dL increase, 95% CI 0.72, 1.64; p=0.70: Table S4). Preoperative-to-24-hour postoperative CSF albumin level changes were not associated with delirium in univariable (Table 2) or multivariable analysis adjusting for preoperative CSF albumin, baseline cognition, age, surgery type, albumin-containing fluid administration, and crystalloid fluid administration (OR=0.91 per 5.8 mg/dL increase, 95% CI 0.55-1.49, p=0.70; Table S5; Figure 3B)

### Albumin Levels and Postoperative Hospital Length of Stay

In both univariable analysis (Table 3) and a negative binomial regression, adjusted for baseline continuous cognitive index, age and surgery group, lower preoperative plasma albumin levels were associated with increased postoperative length of stay (mean ratio [i.e., percent increase] =1.17, 95% CI 1.05-1.31, p=0.004, Table S6). In both univariable analyses (Table 3) and a negative binomial regression also adjusted for 24-hour albumin and crystalloid administration (based on ideal body weight), preoperative plasma albumin level, and delirium incidence, larger preoperative-to-24-hour postoperative plasma albumin decreases were associated with increased postoperative length of stay (mean ratio=1.44, 95% CI 1.26-1.64, p<0.001, Table S7).

**Table 3:** Univariable associations between plasma albumin and CSF albumin, and postoperative Hospital LOS.

|  | Univariable<br>Negative Binomial<br>Regression<br>MR (95% CI) | p-value |
| --- | --- | --- |
| **Plasma Albumin (per 1 SD g/dL decrease)** |  |  |
| Preop (g/dL) | 1.25 (1.12, 1.40) | <0.001 |
| Change (g/dL) | 1.35 (1.18, 1.53) | <0.001 |
| **CSF Albumin (per 1 SD mg/dL increase)** |  |  |
| Preop (mg/dL) | 0.92 (0.79, 1.08) | 0.311 |
| Change (mg/dL) | 1.17 (1.01, 1.34) | 0.034 |

CSF = cerebrospinal fluid, WRS = Wilcoxon Rank Sum, OR = odds ratio, CI = confidence interval.

Preoperative CSF albumin levels were not associated with postoperative hospital length of stay in univariable analysis (OR=0.92, 95% CI 0.79-1.08, p=0.311; Table 3) or in multivariable analysis (Table S8). However, in negative binomial regression adjusted for 24-hour albumin and crystalloid administration (based on ideal body weight), preoperative CSF albumin level, delirium incidence, baseline continuous cognitive index, age and surgery group, larger preoperative-to-24-hour postoperative CSF albumin increases (mean ratio=1.17, 95% CI 1.04-1.32, *p*=0.009, Table S9) were associated with longer postoperative hospital length of stay.

To examine whether the relationships between albumin change and postoperative length of stay differed by delirium status, a stratified analysis was conducted using the same negative binomial regression model. In patients without postoperative delirium, a 1 SD (0.43 g/dL) decrease in preoperative-to-24-hour postoperative plasma albumin was associated with a 39% longer length of stay (IRR=1.39, 95% CI 1.21-1.59, *p*<0.001); in patients with delirium, a 1 SD (0.43 g/dL) decrease in plasma albumin was associated with a 290% increase in length of stay (IRR=2.90, 95% CI 1.78-4.73, *p*<0.001). In patients without delirium, a 1 SD (5.8 mg/dL) increase in preoperative-to-24-hour postoperative CSF albumin was associated with a 14% longer hospital stay (IRR=1.14, 95% CI 1.01-1.29, *p*=0.032), but no significant association was observed in patients with delirium (IRR=1.14, 95% CI 0.68-1.91, *p*=0.620).

## Discussion

In this cohort of 240 older non-cardiac, non-neurologic surgery patients, we found that preoperative-to-24-hour postoperative plasma albumin decreases, but not CSF albumin changes, were independently associated with postoperative delirium. Although prior studies have shown postoperative increases in CPAR,^5, 6^ we found that both plasma and CSF albumin levels decreased 24 hours after surgery. These findings suggest that postoperative increases in CPAR do not always reflect increased CSF albumin levels. Instead, postoperative CPAR increases reflect changes in both plasma and CSF albumin levels and may be substantially influenced by rapid reductions in plasma albumin. Given these rapid postoperative decreases in plasma albumin and the delayed equilibration of albumin between the plasma and CSF compartments, postoperative CPAR should be interpreted cautiously as a standalone marker of acute postoperative blood-brain barrier dysfunction. Future studies should incorporate complementary measures of postoperative BBB dysfunction rather than relying solely on CPAR changes after surgery.

Although our findings suggest that CPAR is an imperfect marker of postoperative BBB dysfunction, they do not rule out a relationship of postoperative BBB dysfunction with postoperative delirium for several reasons. First, because postoperative CPAR changes are moderately correlated with both plasma albumin and CSF albumin changes (Figure S3), postoperative plasma albumin changes alone do not determine postoperative CPAR change. Second, 24-hour postoperative CSF albumin changes are an inadequate measure of BBB permeability, because postoperative decreases in plasma albumin should cause proportional decreases in CSF albumin due to plasma and CSF albumin equilibration.^27^ Thus, postoperative CPAR elevations may still represent BBB dysfunction to some extent, but this depends on the extent of equilibration between compartments. Third, BBB dysfunction 24-48 hours after cardiac surgery has been shown with dynamic contrast enhanced magnetic resonance imaging (DCE-MRI).^28^ Finally, orthopaedic surgery in mice impairs key components of the neurovascular unit, including endothelial cells, pericytes, and astrocytic end-feet.^29^ Thus, our findings suggest that future studies of postoperative BBB dysfunction in humans should employ alternative BBB dysfunction markers, such as early postoperative DCE-MRI,^28^ CSF endothelial injury markers,^30^ or direct neurovascular permeability measurements.^31^

Our findings build on prior studies linking postoperative plasma albumin decreases to delirium after cardiac surgery and joint arthroplasty.^11, 32^ We found a strong association of preoperative-to-24-hour postoperative plasma albumin decreases with delirium, which remained significant after adjustment for preoperative albumin levels, 24-hour perioperative fluid administration, and baseline cognitive function. The mechanisms underlying this relationship remain uncertain. Acute postoperative albumin decreases likely reflect the combined effects of multiple perioperative processes,^33^ including inflammation,^34, 35^ endothelial dysfunction,^36^ fluid shifts,^37^ and redistribution of albumin to the extravascular compartment.^15^ Accordingly, surgery increases transcapillary albumin escape,^15, 38^ whereas albumin synthesis is maintained or increased after abdominal surgery,^15^ and 24-hour renal losses through microalbuminuria are minor (<1 g).^39^ The association between postoperative plasma albumin decreases and delirium persisted despite adjustment for perioperative crystalloid administration and surgery type, suggesting that albumin decline likely reflects fundamental aspects of perioperative stress rather than just dilution from IV fluids or the effects of a particular type of surgery. Furthermore, adjustment for the Charlson comorbidity index did not change the effect estimates or significance of plasma or CSF albumin change models (Table S10). Nevertheless, because we did not directly measure causes of plasma albumin increases, such as inflammation, endothelial injury, or extravascular albumin redistribution, the specific biological processes linking postoperative albumin decreases with delirium cannot be determined here.

Consistent with the idea that postoperative plasma albumin decreases are an integrated biomarker of perioperative stress, we also found that preoperative-to-24-hour postoperative plasma albumin decreases were independently associated with prolonged hospitalisation, even after adjusting for exogenous albumin and crystalloid administration, baseline plasma albumin, delirium incidence, age, and surgery type. This association remained significant in a stratified analysis among patients both with and without delirium, indicating that delirium alone does not fully account for the prolonged hospitalisation associated with postoperative plasma albumin decreases. Thus, postoperative albumin decrease may be an integrated biomarker of surgical stress and heightened inflammatory responses that are associated with increased surgical stress and worse overall recovery, rather than a direct cause of delirium.^40^ Additionally, transcapillary escape of albumin from the intravascular into the interstitial compartment may contribute to tissue oedema;^41^ postoperative fluid accumulation and oedema could, in turn, impede mobility and delay functional recovery.^42^ Since, we did not measure tissue oedema or early postoperative physical function, future studies should evaluate potential factors that underlie worse overall postoperative recovery in older surgical patients with postoperative hypoalbuminemia.

We also observed a modest association between CSF albumin changes and prolonged hospitalisation that was not accompanied by an association with delirium. Because this finding was secondary and the biological significance remains uncertain, it should be interpreted cautiously and requires replication in independent cohorts.

This study has several limitations. First, we cannot fully determine how accurately postoperative CPAR reflects BBB dysfunction because the rate of equilibration between plasma and CSF albumin after surgery is unknown. Rapid postoperative decreases in plasma albumin may therefore alter CPAR independently of BBB permeability, potentially uncoupling postoperative CPAR changes from true changes in BBB function. Additionally, we only measured albumin concentrations before and 24 hours after surgery, preventing characterization of the temporal dynamics of plasma and CSF albumin changes during the early postoperative period. Interindividual variation in the timing of peak postoperative inflammatory/BBB responses and albumin changes may therefore have reduced our statistical power to identify associations between postoperative CSF albumin changes and postoperative delirium risk. Second, the mechanisms linking postoperative albumin decreases with delirium cannot be determined from these data. Although we adjusted for fluid administration and surgery duration, residual confounding by unmeasured markers of perioperative stress remains possible. Because postoperative albumin decreases may reflect the integrated effects of surgical stress, inflammation, endothelial dysfunction, and other perioperative processes, the extent to which albumin serves as a surrogate marker versus a mechanistic mediator of delirium risk cannot be determined from this study. Third, the relatively small number of delirium events (n=31) restricts the number of covariates that can be reliably included in multivariable analyses, raising the possibility of residual confounding. However, we observed a consistent and strong relationship of postoperative plasma albumin decreases with delirium across all univariable and multivariable models. Fourth, although our primary analyses were hypothesis-driven, multiple comparisons were performed across plasma and CSF albumin measures and clinical outcomes. Consequently, the possibility of type I error cannot be excluded, and these findings should be confirmed in independent cohorts. Finally, this was a single-centre study in a predominantly Caucasian, English-speaking, mostly cognitively-intact elective surgery cohort that underwent serial lumbar punctures for research. Thus, the delirium rate observed here may be lower than in those in practice, and these findings may not generalize to higher-risk populations, including patients undergoing emergency surgery, patients with greater medical complexity, or those receiving care outside specialized perioperative research settings.

This study also has several strengths. First, by separately evaluating postoperative changes in plasma and CSF albumin, we were able to test an important assumption underlying interpretation of postoperative CPAR changes, a widely used BBB dysfunction marker. Second, this study leveraged a large, well-phenotyped cohort with paired preoperative and 24-hour postoperative plasma and CSF samples to characterize perioperative albumin dynamics in older patients undergoing non-cardiac, non-neurologic surgery. Finally, our analyses adjusted for important confounders, including baseline cognition and perioperative administration of crystalloids and albumin-containing fluids.

## Conclusions

In 240 older patients undergoing non-cardiac, non-neurologic surgery, preoperative-to-24-hour postoperative decreases in plasma albumin, but not CSF albumin changes, were independently associated with postoperative delirium. Both plasma and CSF albumin levels decreased after surgery, indicating that postoperative CPAR changes reflect alterations in both compartments rather than isolated increases in CSF albumin. The strong association between postoperative plasma albumin decreases and delirium suggests that plasma albumin dynamics may contribute to previously observed relationships between postoperative CPAR increases and delirium. Furthermore, postoperative plasma albumin decreases were associated with prolonged hospital stay, suggesting that plasma albumin decline may serve as an integrated marker of overall postoperative recovery. Overall, these findings highlight important limitations of CPAR as a standalone marker of postoperative blood-brain barrier dysfunction and support the use of complementary measures of BBB integrity in future studies.

## Data Availability

All data produced in the present study are available upon reasonable request to the authors.

## Authors’ Contributions

RH: study design, data acquisition, data interpretation, and manuscript writing; MCW: study design, data analysis and interpretation, and manuscript writing; ERM: study design and data interpretation; NT: data interpretation; JNB: data interpretation; HEW: study conception and design; HJC: study conception and design; AGN: data analysis and interpretation; MKW: study design and data acquisition; JT: manuscript writing and data interpretation; LSD: manuscript writing and data interpretation. KM: data acquisition; ST: data acquisition; MEK: data acquisition; PCB: data interpretation and manuscript drafting; NJT: data interpretation and manuscript drafting; MM: data acquisition and interpretation; EWE: data interpretation; JPM: study conception and data interpretation; MB: study conception, design, data interpretation, and manuscript writing; MJD: study conception, design, data acquisition and interpretation, and manuscript writing.

## Acknowledgements

We thank the patients who participated and the clinical staff who cared for them for making this work possible, and Dr Richard Moon for helpful discussions about postoperative albumin changes.

## Declaration of Interests

Dr. Berger acknowledges additional support from the Duke Anesthesiology Department, the Alzheimer’s Drug Discovery Foundation, NIH T32 GM-08600 (to Dr David S. Warner), UH2-056925 (to HEW), R01-AG073598 (to MB), R01-AG076903 (to MB) and K24-AG103697 (to MB), the Duke Claude D. Pepper Older American Independence Centre (NIH P30-AG028716 to Dr Ken Schmader), the Duke-UNC Alzheimer’s Disease Research Center grant (NIH P30-AG072598 to HEW), and a William L. Young neuroscience research award from the Society for Neuroscience in Anesthesiology and Critical Care (SNACC). JB acknowledges additional funding from R01-HL130443 (to JB, JM), U01-HL088942 (JM, JB), and U01-AG050618 (JB). NT acknowledges support from RF1AG079138 and R01AG057525-06. JPM acknowledges funding from R01HL130443 and R01AG074185. MJD acknowledges funding from K23AG084898, R01AG073598, a Duke-UNC Alzheimer’s Disease Research Center Development Project (from P30-AG072598) Grant, the Duke Anesthesiology Department, and a Merck Investigator Studies Program Grant. HEW acknowledges support from Duke Claude D. Pepper Old American Independence Centre (NIH P30-AG028716) and the Duke-UNC Alzheimer’s Disease Research Center (NIH P30-AG072598).

## Funding

This work was supported by NIH R03AG067976 (MJD), a Foundation for Anesthesia and Education Research GEMSSTAR grant (MJD), NIH K23AG084898 (MJD), an IARS mentored research award (MB), NIH P30-AG072598 (HEW), R01-AG073598 (MB), and NIH grants R03-AG050918 (MB), K76-AG057022 (MB) and K24-AG103697 (MB).

## Declaration of Generative AI and AI-assisted technologies in the writing process

During the preparation of this work the author(s) used Microsoft Copilot to assist with proofreading and language refinement to improve clarity and readability. After using this tool, the authors reviewed and edited the content as needed and take full responsibility for the content of the publication.

## Notes

### Author Declarations

Duke University Health System Institutional Review Board gave ethical approval for this work

