## Supplemental Materials for "Differential Associations of Postoperative Plasma and Cerebrospinal Fluid Albumin Changes with Delirium Following Non-Cardiac Surgery"

**Supplementary Materials**

**Supplementary Methods**

CSF and Blood Collection, Processing and Albumin Assays

**Lumbar Puncture Procedure**

Lumbar punctures were performed by a physician using standard sterile technique under local anesthesia within 1 month before surgery and 24 hours after surgery. Participants were positioned in the seated upright with forward flexion or in the lateral decubitus position when the seated position was not tolerated. Cerebrospinal fluid (CSF) was gently aspirated into a 10-mL polypropylene Luer-Lock syringe and immediately transferred to a prechilled 15-mL polypropylene conical tube (VWR, Radnor, PA) maintained on ice.

**Blood collection**

Whole blood (up to 10 mL) was collected by sterile venipuncture at the same time points as CSF sampling (preoperatively and 24 hours postoperatively) into prechilled K₂EDTA Vacutainer tubes (Becton Dickinson, Franklin Lakes, NJ). Samples were immediately placed on ice following collection.

**Biofluid Processing**

CSF samples were centrifuged at 800 × g for 10 minutes at 4°C to remove cellular components. The acellular supernatant was carefully decanted and aliquoted on ice using low-retention pipette tips into prechilled 1.5-mL polypropylene microcentrifuge tubes (Sarstedt; distributed by VWR). Blood samples were centrifuged at 1370 x g for 15 minutes at 4°C to separate plasma from cellular components. Plasma was subsequently aliquoted into 0.5-mL volumes. All sample handling and aliquoting procedures were performed on ice and completed as rapidly as possible.

**Sample storage**

CSF and plasma aliquots were stored at −80°C within 1 hour of collection. All samples remained frozen at −80°C and were not subjected to any additional freeze-thaw cycles before albumin assay, except for CSF from 5 participants that underwent an additional freeze thaw for sub-aliquoting prior to CSF albumin measurement. To minimize analytical variability, all plasma samples were assayed in 1 large batch, and CSF samples were assayed in three large batches.

**Albumin measurements**

CSF albumin was measured using an immunoturbidimetric microalbumin assay on the UniCel DxC 600 System (Beckman Coulter), using 10 µL of CSF per determination. The standard analytical range was 0.2–97 mg/dL. Calibration was performed using the SYNCHRON Systems microalbumin Calibrator. To ensure accuracy, quality control standards were analyzed daily between batches and with each new calibration and reagent cartridge. The intra-assay coefficient of variation (CV) for CSF albumin measurements was 1.24% (SD, 0.98)

Plasma albumin was measured on a UniCel DxC 600 System using the Beckman Coulter ALB reagent, a bromcresol purple timed-endpoint assay. The assay used 3 µL of plasma and had a manufacturer-specified analytical range of 1.0–7.0 g/dL. The assay was calibrated using the SYNCHRON Systems Multi Calibrator at least every 14 days and following maintenance procedures. To ensure accuracy, quality control standards were analysed prior to the assays, following manufacturer-specified maintenance instructions. Representative manufacturer-reported total imprecision ranges from 1.6% to 2.1% at albumin concentrations of 2.2–5.1 g/dL. The intra-assay coefficient of variation (CV) between duplicate plasma albumin measurements was 0.73% (standard deviation [SD], 1.02).

**Albumin Batch Effects**

CSF albumin assays were completed in 3 main batches. To explore the influence of batch on our results, we included it as a term in our model. This model showed a nonsignificant relationship of preoperative-to-24-hour postoperative CSF albumin changes with postoperative delirium (OR 0.92 95% CI 0.54-1.57, p = 0.75), which was overall similar to our primary model in the manuscript (0.91; 95% CI, 0.55, 1.49; p=0.695). Thus, controlling for the effect of CSF albumin batch did not alter the fact that we observed no significant relationship between preoperative-to-24-hour postoperative CSF albumin changes and postoperative delirium.

Cognitive Testing

Baseline cognitive function was assessed preoperatively using a standardized 14-test neuropsychological battery that has been employed extensively in prior studies of postoperative neurocognitive outcomes. The battery included the Wechsler Test of Adult Reading; the Revised Wechsler Memory Scale and Modified Visual Reproduction Test; the Hopkins Verbal Learning Test; the Randt Short Story Memory Test; Digit Span; Trail Making Tests A and B; Digit Symbol; and the Lafayette Grooved Pegboard Test.

These assessments yielded 14 individual cognitive performance measures that were entered into a factor analysis. Trail Making Test B completion times were truncated at 300 seconds. Scores from Trail Making Tests A and B were negatively log-transformed so that higher scores consistently reflected better cognitive performance, aligning them with the directionality of the remaining measures.

Factor analysis with oblique rotation identified a five-factor solution that accounted for 82% of the variance across test scores. The resulting factors represented five cognitive domains: attention/concentration, structured verbal memory, unstructured verbal memory, visuospatial memory, and executive function. A continuous cognitive index score was calculated as the mean of the five domain scores. This composite measure has been used extensively by our group over the past two decades as a sensitive indicator of baseline cognitive function.^1–35^

**Supplementary Figure 1: Study Timeline**
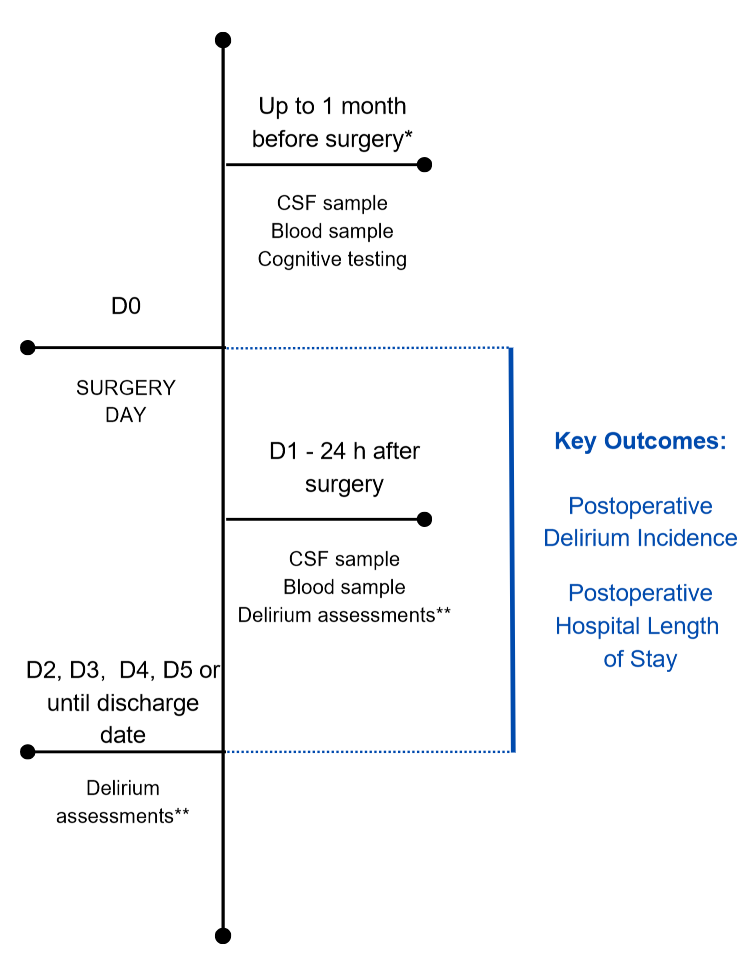

*All cognitive tests, CSF and blood collections were performed up to 1 month before surgery (median [IQR] = 5 [3, 8] days).
**Delirium assessments were conducted once daily in MADCO-PC and twice daily in INTUIT.

**Supplementary Results**

**Supplementary Figure 2:** Both plasma albumin (A) and cerebrospinal fluid albumin levels (B) decrease from before to 24-hours after surgery (n=240).

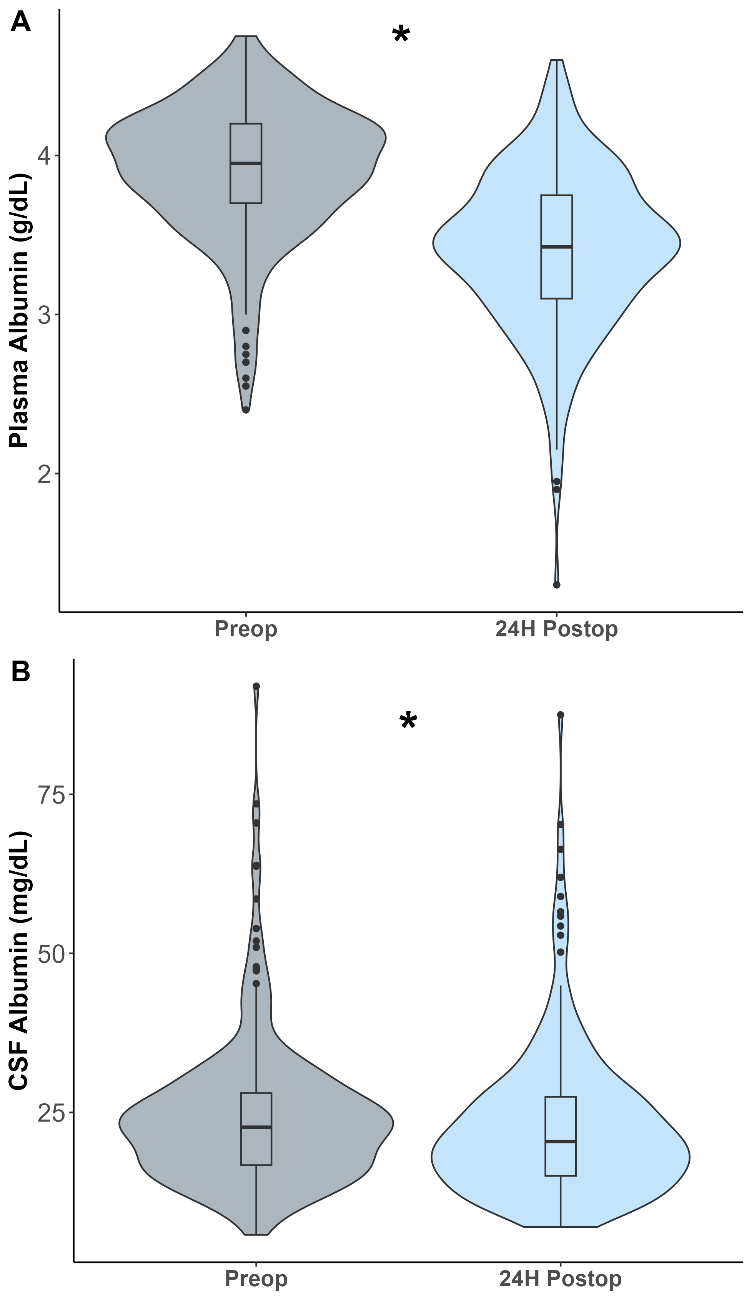

*P<0.001 in Wilcoxon Rank Sum.

**Supplementary Figure 3:** Correlation of preoperative-to-24-hour postoperative (A) plasma albumin change (p<0.001) and (B) CSF albumin change (p<0.001) with preoperative-to-24-hour postoperative CSF: plasma albumin ratio (CPAR) change,
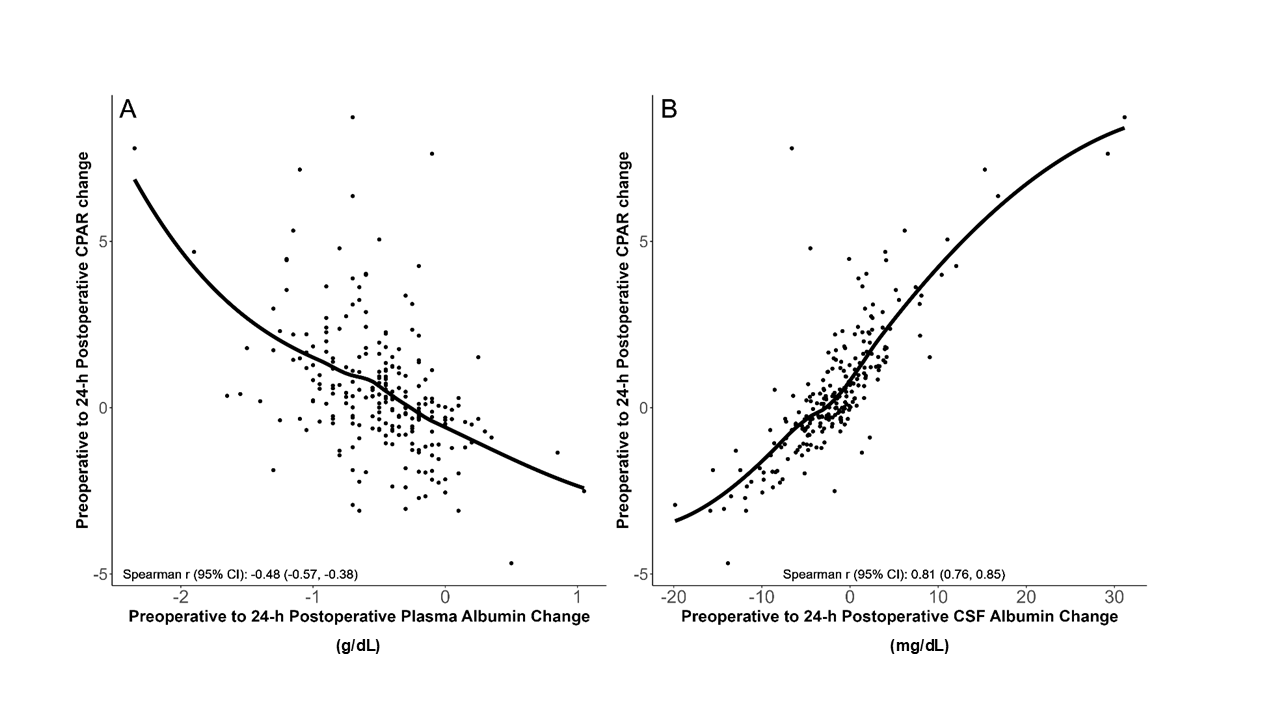
with LOESS fit lines.

CPAR = CSF: plasma albumin ratio, CSF = cerebrospinal fluid, CI = confidence intervals.

**Supplementary Table 1:** Logistic regression examining effect of preoperative plasma albumin levels on postoperative delirium risk.

| **Effect** | **OR (95% CI)** | **p-value** |
| --- | --- | --- |
| Preoperative Plasma Albumin (per 0.42 g/dL) decrease) | 0.88 (0.57, 1.37) | 0.576 |
| Baseline Continuous Cognitive Index (per 0.72 decrease) | 2.88 (1.80, 4.63) | <0.001 |
| Age (per year) | 0.99 (0.91, 1.06) | 0.704 |
| Surgery Group |  |  |
| Open Intra-Abdominal | 2.81 (0.91, 8.67) | 0.072 |
| Orthopaedics | 0.81 (0.26, 2.56) | 0.719 |
| Thoracic | 0.97 (0.25, 3.77) | 0.965 |
| General abdominal, urologic, plastic, gynaecologic, ENT | Ref | Ref |
| *Model fit diagnostics: Akaike Information Criterion = 160.753, area under the receiver operating characteristic curve = 0.79 (95% CI: 0.70-0.88), rescaled R² = 0.221, and the Hosmer-Lemeshow goodness-of-fit test (p = 0.528).* | | |

OR = Odds ratio, 95% CI = 95% confidence intervals.

**Supplementary Table 2:** Logistic regression examining effect of preoperative-to-24-hour postoperative plasma albumin change on postoperative delirium risk.

| **Effect** | **OR (95% CI)** | **p-value** |
| --- | --- | --- |
| Preoperative-to-24-hour Postoperative Plasma Albumin change (per 1 SD (0.43 g/dL) decrease) | 2.09 (1.25, 3.49) | 0.005 |
| Preoperative Plasma Albumin (per 10.42 g/dL lower) | 1.03 (0.62, 1.69) | 0.924 |
| Baseline Continuous Cognitive Index (per 0.72 lower) | 3.52 (2.03, 6.11) | <0.001 |
| Age (per year) | 1.00 (0.93, 1.09) | 0.935 |
| Surgery Group |  |  |
| Open Intra-Abdominal | 1.82 (0.44, 7.61) | 0.412 |
| Orthopaedics | 0.85 (0.22, 3.29) | 0.817 |
| Thoracic | 0.72 (0.15, 3.52) | 0.688 |
| General Abdominal, Urologic, Plastic, Gynaecologic, ENT | Ref | Ref |
| Albumin 5% (10 mL per IBW kg from surgery start to 24 hours later) | 0.68 (0.29, 1.65) | 0.397 |
| Crystalloid (10 mL per IBW kg from surgery start to 24 hours later) | 1.46 (1.09, 1.96) | 0.012 |
| *Model fit diagnostics: Akaike Information Criterion = 145.77, area under the receiver operating characteristic curve = 0.86 (95% CI: 0.79-0.93), rescaled R² = 0.364, and the Hosmer-Lemeshow goodness-of-fit test (p = 0.830).* | | |

OR = Odds ratio, 95% CI = 95% confidence intervals, ENT = ear nose and throat, IBW = ideal body weight.

**Supplementary Table 3:** Associations between plasma and cerebrospinal fluid albumin with delirium first detected after postoperative day 1 (n=16).

|  | **Univariable Logistic Regression**  **OR (95% CI)** | **p-value** | **Multivariable Logistic Regression**  **OR (95% CI)** | **p-value** |
| --- | --- | --- | --- | --- |
| **Plasma Albumin**  **(per 1 SD decline)** |  |  |  |  |
| Preop (g/dL) | 1.24 (0.77, 2.01) | 0.376 | 1.19 (0.71, 2.01) | 0.513 |
| Change (g/dL) | 1.84 (1.14, 2.98) | 0.013 | 2.13 (1.21, 3.75) | 0.009 |
| **CSF Albumin**  (**per 1 SD increase)** |  |  |  |  |
| Preop (mg/dL) | 1.03 (0.60, 1.77) | 0.919 | 1.01 (0.54, 1.89) | 0.987 |
| Change (mg/dL) | 1.32 (0.74, 2.36) | 0.344 | 1.33 (0.71, 2.49) | 0.367 |

OR= odds ratio, 95% CI = 95% confidence intervals, SD = standard deviation, CCI = continuous cognitive index.

**Supplementary Table 4:** Logistic regression examining effects of preoperative CSF albumin levels on postoperative delirium risk.

| **Effect** | **OR (95% CI)** | **p-value** |
| --- | --- | --- |
| CSF Albumin at Baseline (per 11.6 mg/dL increase) | 1.09 (0.72, 1.64) | 0.696 |
| Baseline Continuous Cognitive Index (per 0.72 lower) | 2.85 (1.78, 4.55) | <0.001 |
| Age (per year) | 0.99 (0.91, 1.06) | 0.714 |
| Surgery Group |  |  |
| Open Intra-Abdominal | 2.81 (0.91, 8.67) | 0.072 |
| Orthopaedics | 0.81 (0.25, 2.56) | 0.713 |
| Thoracic | 0.91 (0.24, 3.54) | 0.894 |
| General Abdominal, Urologic, Plastic, Gynaecologic, ENT | Ref | Ref |
| *Model fit diagnostics: Akaike Information Criterion: 160.924, area under the receiver operating characteristic curve = 0.78, 95% CI: 0.69-0.87, rescaled R² = 0.220, and the Hosmer-Lemeshow goodness-of-fit test (p = 0.505).* | | |

OR = Odds ratio, 95% CI = 95% confidence intervals, CCI = continuous cognitive index, ENT = ear nose and throat.

**Supplementary Table 5:** Logistic regression examining effect of preoperative-to-24-hour postoperative CSF albumin change on postoperative delirium risk.

| **Effect** | **OR (95% CI)** | **p-value** |
| --- | --- | --- |
| Preoperative-to-24-hour CSF albumin change (per 5.8 mg/dL increase) | 0.91 (0.55, 1.49) | 0.695 |
| Preoperative CSF Albumin (per 11.6 mg/dL increase) | 1.13 (0.75, 1.68) | 0.567 |
| Baseline Continuous Cognitive Index (per 0.72 lower) | 3.24 (1.94, 5.42) | <0.001 |
| Age (per year) | 0.99 (0.92, 1.08) | 0.871 |
| Surgery Group |  |  |
| Open Intra-Abdominal | 3.20 (0.84, 12.21) | 0.088 |
| Orthopaedics | 1.53 (0.43, 5.43) | 0.514 |
| Thoracic | 1.25 (0.30, 5.22) | 0.765 |
| General abdominal, urologic, plastic, gynaecologic, ENT | Ref | Ref |
| Albumin 5% (10ml per IBW kg from surgery start to 24 hours later) | 0.54 (0.23, 1.26) | 0.156 |
| Crystalloid (10 ml per IBW kg from surgery start to 24 hours later) | 1.60 (1.22, 2.10) | 0.001 |
| *Model fit diagnostics: Akaike Information Criterion = 154.829, area under the receiver operating characteristic curve = 0.82, 95% CI: 0.74-0.91, rescaled R² = 0.304, and the Hosmer-Lemeshow goodness-of-fit test (p = 0.494).* | | |

OR = Odds ratio, 95% CI = 95% confidence intervals, ENT = ear nose and throat, IBW = ideal body weight.

**Supplementary Table 6:** Negative binomial regression examining effect of preoperative plasma albumin levels on postoperative length of stay (in days).

| **Effect** | **MR (95% CI)** | **p-value** |
| --- | --- | --- |
| Preoperative Plasma Albumin (per 0.42 g/dL lower) | 1.17 (1.05, 1.31) | 0.004 |
| Baseline Continuous Cognitive Index (per.72) lower) | 1.15 (1.00, 1.32) | 0.047 |
| Age (per year) | 1.01 (0.99, 1.03) | 0.316 |
| Surgery Group |  |  |
| Open Intra-Abdominal | 2.44 (1.69, 3.52) | <0.001 |
| Orthopaedics | 0.90 (0.65, 1.24) | 0.510 |
| Thoracic | 2.48 (1.74, 3.54) | <0.001 |
| General Abdominal, Urologic, Plastic, Gynaecologic, ENT | Ref | Ref |

MR = Mean Ratio from negative binomial regression, 95% CI = 95% confidence intervals, SD = standard deviation, ENT = ear nose and throat.

**Supplementary Table 7:** Negative binomial regression examining effect of preoperative-to-24-hour postoperative plasma albumin change on postoperative hospital length of stay (in days).

| **Effect** | **MR (95% CI)** | **p-value** |
| --- | --- | --- |
| Preoperative-to-24-hour Postoperative Plasma Albumin change (per 0.43 g/dL decrease) | 1.44 (1.26, 1.64) | <0.001 |
| Preoperative Plasma Albumin (per 0.42 g/dL decrease) | 1.31 (1.17, 1.46) | <0.001 |
| Baseline Continuous Cognitive Index (per 0.72 lower) | 1.04 (0.90, 1.20) | 0.580 |
| Age (per year) | 1.01 (0.99, 1.03) | 0.345 |
| Surgery Group |  |  |
| Open Intra-Abdominal | 1.26 (0.86, 1.84) | 0.243 |
| Orthopaedics | 0.91 (0.66, 1.25) | 0.569 |
| Thoracic | 2.21 (1.60, 3.06) | <0.001 |
| General Abdominal, Urologic, Plastic, Gynaecologic, ENT | Ref | Ref |
| Albumin 5% (10 mL per IBW kg from surgery start to 24 hours later) | 1.54 (1.21, 1.94) | <0.001 |
| Crystalloid (10 mL per IBW kg from surgery start to 24 hours later) | 1.05 (0.97, 1.13) | 0.208 |
| Delirium | 1.62 (1.14, 2.29) | 0.007 |

MR = Mean Ratio from negative binomial regression, 95% CI = 95% confidence intervals, SD = standard deviation, CCI = continuous cognitive index, ENT = ear, nose and throat, IBW = ideal body weight.

**Supplementary Table 8:** Negative binomial regression examining effect of preoperative CSF albumin levels on postoperative length of stay (in days).

| **Effect** | **MR (95% CI)** | **p-value** |
| --- | --- | --- |
| Preoperative CSF Albumin (per 11.6 mg/dL increase) | 0.95 (0.83, 1.09) | 0.476 |
| Baseline Continuous Cognitive Index (per 0.72 decrease) | 1.20 (1.04, 1.38) | 0.010 |
| Age (per year) | 1.01 (0.99, 1.03) | 0.322 |
| Surgery Group |  |  |
| Open Intra-Abdominal | 2.28 (1.58, 3.30) | <0.001 |
| Orthopaedics | 0.85 (0.62, 1.18) | 0.337 |
| Thoracic | 2.67 (1.87, 3.82) | <0.001 |
| General abdominal, Urologic, Plastic, Gynaecologic, ENT | Ref | Ref |

MR = Mean Ratio from negative binomial regression, 95% CI = 95% confidence intervals, SD = standard deviation, ENT = ear, nose and throat.

**Supplementary Table 9:** Negative binomial regression model for preoperative-to-24-hour CSF albumin change and postoperative hospital length of stay (in days).

| **Effect** | **MR (95% CI)** | **p-value** |
| --- | --- | --- |
| Preoperative-to-24-hour Postoperative CSF Albumin (per 5.8 mg/dL increase) | 1.17 (1.04, 1.32) | 0.009 |
| Preoperative CSF Albumin (per 11.6 mg/dL higher) | 1.01 (0.89, 1.16) | 0.855 |
| Baseline Continuous Cognitive Index (per 0.72 lower) | 1.09 (0.94, 1.26) | 0.239 |
| Age (per year) | 1.01 (0.99, 1.03) | 0.342 |
| Surgery Group |  |  |
| Open Intra-Abdominal | 1.58 (1.07, 2.34) | 0.022 |
| Orthopaedics | 1.02 (0.74, 1.42) | 0.884 |
| Thoracic | 2.83 (2.03, 3.96) | <0.001 |
| General abdominal, urologic, plastic, gynaecologic, ENT | Ref | Ref |
| Albumin 5% (10 mL per IBW kg from surgery start to 24 hours later) | 1.31 (1.02, 1.67) | 0.032 |
| Crystalloid (10 mL per IBW kg from surgery start to 24 hours later) | 1.08 (1.00, 1.17) | 0.051 |
| Delirium | 2.01 (1.41, 2.87) | <0.001 |

MR = Mean Ratio from negative binomial regression, 95% CI = 95% confidence intervals, SD = standard deviation, ENT = ear, nose and throat, IBW = ideal body weight.

### **Supplementary Table 10:** Sensitivity Analysis: Associations of Perioperative Albumin Changes With Postoperative Outcomes After Multivariable Adjustment for Age, Surgery Group, Albumin 5% Administration, Crystalloid Administration, and Charlson Comorbidity Index

|  | **Postoperative Delirium Risk  OR (95% CI)** | **p-value** | **Hospital Length of Stay  MR (95% CI)** | **p-value** |
| --- | --- | --- | --- | --- |
| **Plasma Albumin (per 0.43 g/dL decrease)** |  |  |  |  |
| Effect of preop-to-24-hour postop change | 2.12 (1.26, 3.54) | 0.0045 | 1.43 (1.26, 1.64) | <0.001 |
| **CSF Albumin (per 5.8 mg/dL increase)** |  |  |  |  |
| Effect of preop-to-24-hour postop change | 0.98 (0.90, 1.07) | 0.664 | 1.17 (1.04, 1.32) | 0.009 |

OR = odds ratio; MR = mean ratio. 95% CI = 95% confidence intervals. Odds ratios were estimated using logistic regression models for delirium. Mean ratios were estimated using negative binomial regression models for hospital length of stay.
